# Validation of the 4C Deterioration Model for COVID-19 in a UK teaching hospital during Wave 2

**DOI:** 10.1101/2021.06.22.21259331

**Authors:** Sarah L. Cowan, Martin Wiegand, Jacobus Preller, Robert J. B. Goudie

## Abstract

The 4C Deterioration model was developed and validated on data collected in UK hospitals until August 26, 2020, but has not yet been validated in the presence of SARS-CoV-2 variants and novel treatment regimens that have emerged subsequently. In this first validation study of the 4C Deterioration model on patients admitted between August 27, 2020 and April 16, 2021 we found, despite a slightly overestimation of risk, that the discrimination (area under the curve 0.75, 95% CI 0.71-0.78) and calibration of the model remained consistent with the development study, strengthening the evidence for adopting this model into clinical practice.

---

The 4C Deterioration model is a point-of-admission tool for predicting in-hospital clinical deterioration in patients with COVID-19.^1^ It was developed and validated using data from the UK first wave until August 26, 2020. The vast majority of point-of-admission models proposed for risk stratification in COVID-19 suffer from poor calibration.^2^ In contrast the 4C Deterioration model was shown to be well-calibrated and have good discriminative characteristics.^1^

Since the model was developed, treatment of COVID-19 has evolved, including the use of corticosteroids as standard of care in hypoxemic respiratory failure, and new variants of the have virus emerged.^3,4,5^ Such changes can lead to declining model performance over time.^6^ Temporal validation during the second wave of the pandemic is therefore important to assess whether the discrimination and calibration of the model has been maintained.

Here we present the first external validation of the 4C Deterioration Model using data from the UK second wave.

## Methods

All adult patients admitted to Cambridge University Hospitals between August 27, 2020 and April 16, 2021 who tested positive for SARS-CoV-2 were included in the validation cohort. Diagnostic testing used either a real-time reverse transcription polymerase chain reaction (RT-PCR) of the RdRp gene from a nasopharyngeal swab, or the SAMBA II point-of-care test used at the hospital.^7^ Readmissions and inter-hospital transfers were excluded. Data were extracted from the electronic health record system (Epic) on May 14, 2021, meaning at least 28 days follow up were available for all included patients. The data were analysed retrospectively in R 3.6.3.

All patients were treated as per detailed local guidance in use in the hospital at the time.

As in the development study, the outcome was in-hospital deterioration (commencement of ventilatory support, critical care admission, or death); cases tested more than seven days after admission were considered nosocomial; and patients who remained in hospital but had not deteriorated by the time of data extraction were classed as not deteriorating.

The 4C Deterioration model uses a combination of demographic factors (age and sex), blood tests (C-reactive protein, urea, lymphocytes), observations (respiratory rate, oxygen saturation (SpO2), Glasgow Coma Scale), requirement for supplemental oxygen, whether the infection was hospital-acquired, and the presence of lung infiltrates on radiographic chest imaging.^1^

To calculate the risk score we used only results and observations recorded within 24 hours of admission, or within 24 hours of the time of first positive test for nosocomial cases. The development study accounted for missing values in their data using multiple imputation but did not report the imputation model parameters used, meaning that this approach cannot be used either in validation or in clinical practice. Missing values in the validation data were instead median imputed from the development dataset.^8^ This avoids the potential bias that would be introduced if only patients with a complete set of observations, blood results and imaging were included in the validation, as the presence or absence of observations or tests may in itself reflect clinician assessment of the severity of disease.

To assess the discriminative performance of the proposed model we calculate the Area under the Receiver-Operating Curve (AUROC), where a value of 1 represents perfect discrimination and 0·5 discrimination no better than random chance. Additionally we stratify the AUROC by month of patient admission, to investigate performance over time. We also calculate the Area under the Precision-Recall Curve (AUPRC; also known as the curve of positive predictive value (PPV) against sensitivity; true positives/(all positives) - true positives/(true positives + false negatives)), which measures discrimination relative to the observed incidence; and the number needed to evaluate (NNE = 1/PPV), defined as the number of patients predicted to deteriorate for every one additional correctly-detected deterioration, which is a measure of clinical burden.^9^ We assess model calibration using calibration-in-the-large and the calibration slope.^10^ We also visualise the calibration of the model through the observed incidence in each decile of predicted deterioration probability.

The study was approved by a UK Health Research Authority ethics committee (20/WM/0125). Patient consent was waived because the de-identified data presented here were collected during routine clinical practice; there was no requirement for informed consent.

## Results

950 patients were included. Compared to the development study, patients were younger (median 70 years vs 75) and nosocomial infections were slightly more common (11·2% vs 9·9%). Other parameters were similar (Table 1). Missingness was greatest for urea (21·1% missing), radiology (15·9%), C-reactive protein (11·2%) and lymphocyte count (9·3%), all lower than in the development study.

**Table 1:** Patient baseline characteristics and distribution of parameters.

|  | **Our data** | **ISARIC 4C development study** |
| --- | --- | --- |
| Number of patients | 950 | 74 944 |
| Male, n (%) | 498 (52.4%) | 41 993 (56.1%) |
| Age, median [IQR] | 70 [53, 82] | 75 [60, 84] |
| Nosocomial infection, n (%) | 106 (11.2%) | 7320 (9.9%) |
| Glasgow Coma Scale, median [IQR] | 15 [15, 15] | 15 [15, 15] |
| Respiratory Rate, breaths per minute, median [IQR] | 19 [17, 23] | 20 [18, 26] |
| Oxygen saturation, %, median [IQR] | 96 [94, 97] | 95 [92, 97] |
| Room air, n (%) | 573 (60.3%) | 48574 (69.4%) |
| Urea, mmol/L, median [IQR] | 6.2 [4.5, 9.3] | 7 [5, 11] |
| C-reactive protein, mg/L, median [IQR] | 57 [22, 113] | 80 [33, 154] |
| Lymphocyte count, x 10<sup>9</sup>/L, median [IQR] | 0.85 [0.59, 1.21] | 0.9 [0.6, 1.3] |
| Radiographic infiltrates / number of patients with radiology result available | 497/807 (61.6%) | 29 579 / 47 749 (61.9%) |
| **Outcomes\*** |  |  |
| Ventilatory support or critical care admission, n (%) | 182 (19.2%) | 15 039 (20.1%) |
| Death, n (%) | 99 (10.4%) | 16 885 (22.5%) |
| No deterioration, n (%) | 669 (70.4%) | 42 024 (56.1%) |
| Missing, n (%) | 0 (0.0%) | 996 (1.3%) |
| **Model performance** |  |  |
| AUROC [95% CI] | 0.75 [0.71 to 0.78] | 0.77 [0.76 to 0.78] |
| Calibration-in-the-large [95% CI] | -0.26 [-0.42 to -0.11] | 0.00 [-0.05 to 0.05] |
| Calibration slope [95% CI] | 1.00 [0.83 to 1.18] | 0.96 [0.91 to 1.01] |

\* Outcomes given here are the first point at which patients fulfil the composite endpoint for deterioration.

In-hospital deterioration occurred in 281 (29·6%) patients, compared to 42·6% in the development study. The lower risk of deterioration in this cohort may reflect differences in the patient population of the study hospital, or could represent improvements in treatment over the course of the pandemic.

Figure 1 shows the performance metrics for the median-imputed data set. AUROC was 0·75 [95% CI 0·71 to 0·78]; calibration-in-the-large was -0·26 [-0·42 to -0·11], indicating overprediction of risk; and the calibration slope was 1·00 [0·83 to 1·18]. The NNE remains below 3·5 over the entire range of sensitivity, indicating that the clinical burden of use of the score is reasonable. The assessment of AUROC by month of admission (eFigure 1) only revealed a slight decrease in the discriminative performance during the winter, when bed occupancy was reaching its peak.

**Figure 1.**
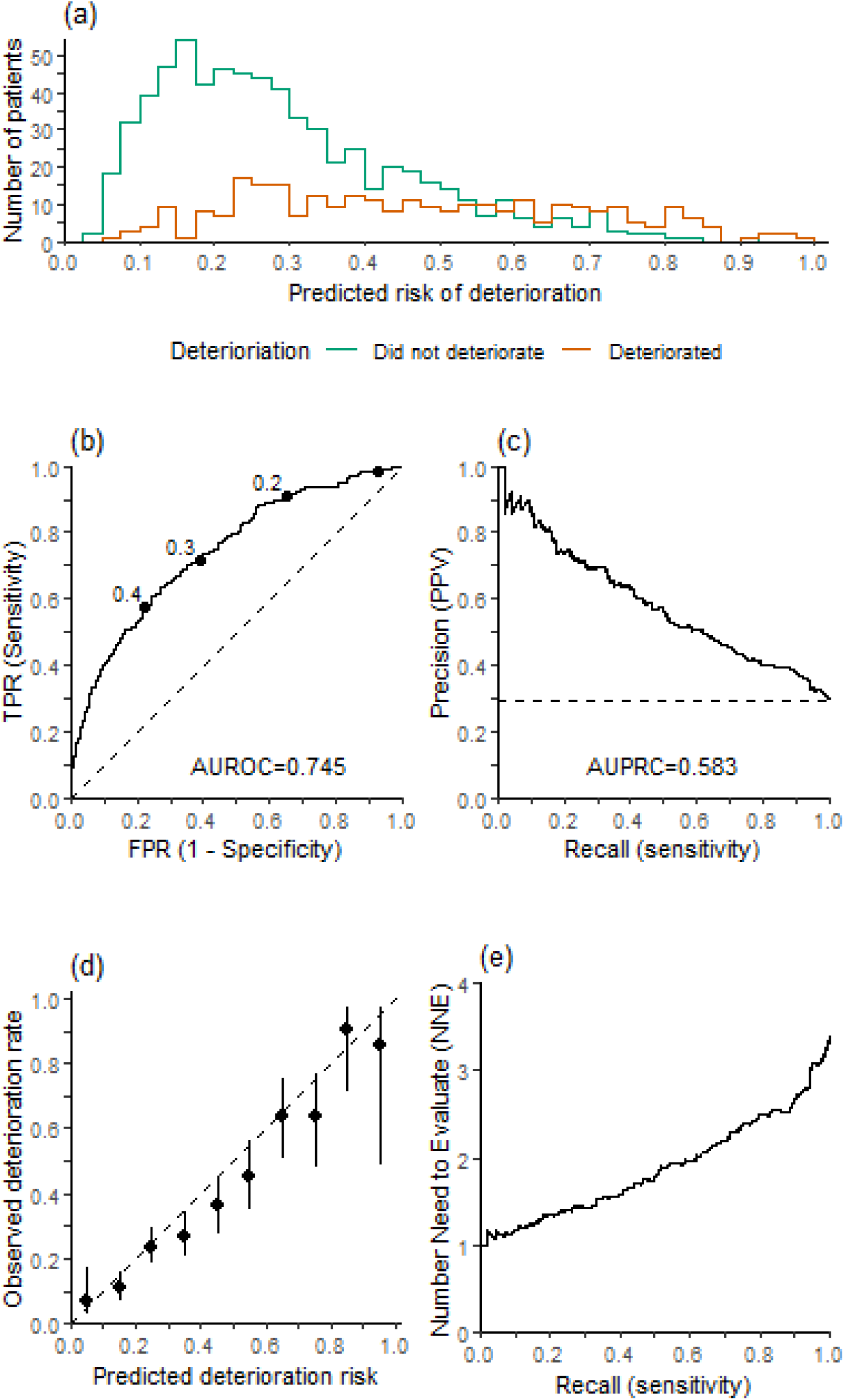
(a) Histogram of predicted risk of clinical deterioration; (b) Receiver Operator Characteristic plot, with labels indicating the corresponding cutoff and the dashed line indicating the line of no discrimination; (c) Precision-Recall plot, with the 29·6% observed deterioration incidence indicated by the dashed line; (d) Calibration plot (with 95% CI), by tenths of predicted risk, with the dashed line indicating perfect calibration. AUROC = Area under the Receiver Operator Curve; TPR = true positive rate; FPR = false positive rate; AUPRC = Area under the Precision Recall Curve; PPV = positive predictive value. (e) Number needed to evaluate (NNE) by sensitivity (recall).

Median imputation proved to be a viable approach to missing data, as the model’s performance was not adversely affected by the imputed values. Additionally this suggests that the model can be applied more widely, as many patients had missing values in at least one predictor: 384 (40.4%) patients in our study had at least one missing observation or result; compared to at least 36.3% (missing chest imaging alone; overall missingness unreported) in the development study. Performance was similar when patients with any missingness were excluded (AUROC 0·78 [0·74 to 0·82]; calibration-in-the-large -0·28 [-0·48 to -0·09]; calibration slope 1·09 [0·88 to 1·31]).

## Conclusion

Despite slight overestimation of risk, discrimination and calibration remained consistent with the development study demonstrating robustness to the presence of novel variants and changes in treatment over time.

## Data Availability

The de-identified data that support the findings of this study are available from Cambridge
University Hospitals but restrictions apply to the availability of these data, which were used
under license for the current study, and so are not publicly available. Data are however
available from the authors upon reasonable request with permission of Cambridge University
Hospitals.

## Acknowledgements

Martin Wiegand was funded by the NIHR Cambridge Biomedical Research Centre (BRC-1215-20014). Robert J. B. Goudie was funded by the UKRI Medical Research Council [programme code MC_UU_00002/2] and supported by the NIHR Cambridge Biomedical Research Centre (BRC-1215-20014). The Clinical Informatics data extraction was funded by the Cancer Research UK Cambridge Centre and conducted by Vince Taylor. The views expressed are those of the authors and not necessarily those of the NHS, the NIHR or the Department of Health and Social Care.

The funders had no role in the design and conduct of the study; collection, management, analysis, and interpretation of the data; preparation, review, or approval of the manuscript; and decision to submit the manuscript for publication.

## Authorship Contribution Statement

SLC, JP and RJBG conceived the design of the work. MW, SLC and RJBG prepared the data and conducted the statistical analysis. All authors contributed to the drafting and the revision of the manuscript. The final version of the manuscript has been approved by all authors, and are accountable for the accuracy and integrity of the work presented.

## Supplementary Material

**eFigure 1.**
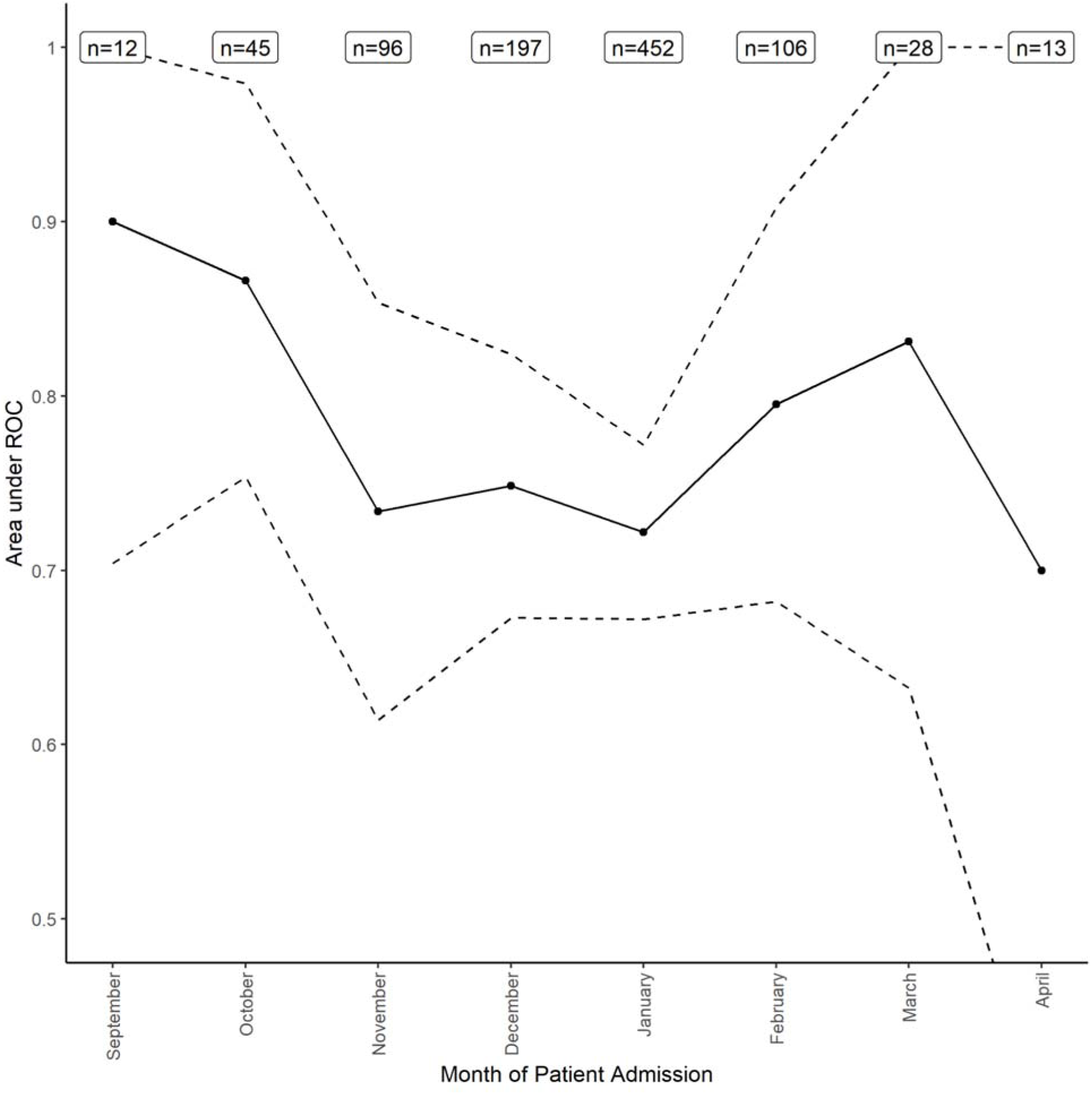
Area under the ROC plot, by admission date. The dashed line indicates the 95% CI.

## References

1 Gupta RK, Harrison EM, Ho A et al. Development and validation of the ISARIC 4C Deterioration model for adults hospitalised with COVID-19: a prospective cohort study. Lancet Respir Med. 2021; 2(4): e592–359. https://doi.org/10.1016/S2213-2600(20)30559-2

2 Wynants L, Van Calster B, Collins GS et al. Prediction models for diagnosis and prognosis of COVID-19: Systematic review and critical appraisal. BMJ. 2020; 369: m1328. https://doi.org/10.1136/bmj.m1328

3 Volz E, Mishra S, Chand M et al. Assessing transmissibility of SARS-CoV-2 lineage B.1.1.7 in England. Nature. 2021; 593: 266–269. https://doi.org/10.1038/s41586-021-03470-x

4 Davies NG, Jarvis CI, CMMID COVID-19 Working Group. et al. Increased mortality in community-tested cases of SARS-CoV-2 lineage B.1.1.7. Nature 593, 270–274 (2021). https://doi.org/10.1038/s41586-021-03426-1

5 The RECOVERY Collaborative Group. Dexamethasone in Hospitalized Patients with Covid-19. N Engl J Med. 2021; 384: 693–704. https://doi.org/10.1056/NEJMoa2021436

6 Nashef SAM, Roques F, Sharples LD et al. EuroSCORE II. Eur J Cardiothorac Surg. 2012; 41(4): 734–44. https://doi.org/10.1093/ejcts/ezs043

7 Assennato SM, Ritchie AV, Nadala C, et al. Performance Evaluation of the SAMBA II SARS-CoV-2 Test for Point-of-Care Detection of SARS-CoV-2. J Clin Microbiol. 2020; 59(1): e01262–20. https://doi.org/10.1128/JCM.01262-20

8 Nijman SW, Groenhof TK, Hoogland J et al. Real-time imputation of missing predictor values improved the application of prediction models in daily practice. J Clin Epidemiol. 2021; 134: 22–34. https://doi.org/10.1016/j.jclinepi.2021.01.003

9 Romero-Brufau S, Huddleston JM, Escobar GJ et al. Why the C-statistic is not informative to evaluate early warning scores and what metrics to use. Crit Care. 2015; 19: 285. https://doi.org/10.1186/s13054-015-0999-1

10 Steyerberg EW. Clinical Prediction Models: A Practical Approach To Development, Validation and Updating. Springer, Cham, 2019.

